# Impact of Chronic Kidney Disease on Clinical Outcomes after Percutaneous Coronary Intervention with Drug-Coated Balloons

**DOI:** 10.1101/2024.06.20.24309055

**Authors:** Tetsuya Takahashi, Tetsu Watanabe, Mashu Toyoshima, Wataru Katawaki, Taku Toshima, Yu Kumagai, Tamon Yamanaka, Masafumi Watanabe

## Abstract

**Background:** Drug coated balloon (DCB) is an emerging treatment technology for percutaneous coronary intervention (PCI). However, the prognostic factors of PCI with DCB remain fully determined. Chronic kidney disease (CKD) is an independent predictor of adverse outcomes in patients with coronary artery disease (CAD) who underwent PCI. The aim of the present study was to clarify the impact of CKD on prognosis in CAD patients who underwent PCI with DCB.

**Methods:** We enrolled 252 consecutive patients with CAD who underwent PCI with DCB from 2015 to 2023. The endpoints of this study were composite events including all-cause death, myocardial infarction, target vessel revascularization, stroke, and major bleeding.

**Results:** The prevalence rate of CKD was 48%. Patients with CKD were older and had higher prevalence of hypertension and diabetes mellitus than those without. Kaplan-Meier analysis revealed a significantly higher composite event rate in patients with CKD (log-rank test, P = 0.003). In the multivariate Cox proportional hazards analysis, CKD was independently associated with composite events after adjusting for confounding factors (adjusted hazard ratio 1.920, 95% confidence intervals 1.154-3.197, P = 0.012), mainly driven by all-cause deaths.

**Conclusion:** CKD was associated with unfavorable outcomes in CAD patients who underwent PCI with DCB.

## Introduction

Drug-coated balloons (DCBs) have been established as a novel treatment option of percutaneous coronary intervention (PCI) ^1, 2, 3^. The previous study has demonstrated that DCB treatment has been noninferior compared to conventional drug uncoated balloon angioplasty for the treatment of in-stent restenosis ^1^. DCB has also shown comparable clinical outcomes compared to drug eluting stent (DES) in patients with small-vessel coronary artery disease (CAD) ^2, 3^. Furthermore, recent studies have demonstrated the favorable clinical effect of DCB in CAD patients with multivessel, de novo large vessel, and long diffuse vessel diseases^4, 5, 6^. However, the prognostic impact of DCB on clinical outcomes in patients with CAD has not been adequately addressed although there are several favorable clinical data in the setting of comparison between DCB and DES.

Chronic kidney disease (CKD) is reportedly observed in approximately 10% of the general population ^7^. The prevalence rate of CKD in patients with CAD has been increasing with aging population ^8, 9^. The previous study demonstrated that CAD severity was associated with the CKD ^10^. The presence of CKD has been associated with unfavorable prognosis in patients with CAD ^11^. Therefore, CKD has been recognized as one of the important predictor for cardiovascular prognosis in the era of DES ^12^. However, the association between CKD and DCB remains unclear.

The aim of the present study was to clarify the prognostic impact of CKD on prognosis in CAD patients who underwent PCI using DCB.

## Methods

### Study population

This is a prospective, single-center, observational study. We enrolled 288 consecutive CAD patients who underwent PCI with DCB between January 2015 and December 2023. We excluded 36 patients with missing clinical data after PCI. The remaining 252 patients were included in this study.

Acute coronary syndrome (ACS) and chronic coronary syndrome (CCS) were diagnosed based on the current guidelines, respectively ^13, 14^.

Angiographical coronary stenosis was assessed according to the American College of Cardiology / American Heart Association / Society for Cardiovascular Angiography and Interventions (ACC / AHA / SCAI) ^15^. Significant coronary stenosis was defined as a visually estimated diameter stenosis severity of ≥ 70% for non-left main disease and ≥ 50% for left main disease.

PCI was performed based on contemporary standard techniques and current guidelines ^16, 17, 18^.

Demographic and clinical data including age, sex, smoking, atrial fibrillation (AF), family history of CAD, previous myocardial infarction (MI), previous PCI, previous coronary artery bypass graft (CABG), previous stroke, chronic heart failure (CHF), clinical presentation of peripheral artery disease (PAD), abdominal aortic aneurysm (AAA), chronic obstructive pulmonary disease (COPD), malignant tumor, ACS and CCS, and medications at discharge were collected from patients’ medical records and through interviews. Body mass index (BMI) was calculated based on the patient’s weight and height measured during hospitalization. Diagnoses of hypertension, diabetes mellitus, and dyslipidemia were established based on medical records or a history of related medical therapy. Transthoracic echocardiography was performed by physicians.

Informed consent was obtained from all patients prior to participation. All procedures were performed in accordance with the Declaration of Helsinki. The study was approved by the Institutional Ethics Committee of Yamagata University School of Medicine (Yamagata University, 2020-344).

### Assessment of renal function

The eGFR was calculated using the Modification of Diet in Renal Disease equation with the Japanese coefficient ^19^. Venous blood samples were obtained early in the morning, within 24 h after admission. Serum creatinine levels were measured using routine laboratory methods. Chronic kidney disease (CKD) was defined as reduced eGFR (< 60 mL/min/1.73m^2^) according to Kidney Disease Outcomes Quality Initiative clinical guidelines ^20^.

### Endpoints and follow-up

Patients were prospectively followed up for a median duration of 492 days (interquartile range 248 -1532 days). Clinical follow-up data were obtained from outpatient record reviews and telephone interviews. The primary endpoint of the present study was composite events including all-cause death, myocardial infarction, target vessel revascularization, stroke, and major bleeding (Bleeding Academic Research Consortium [BARC] type 3-5) ^21^. The secondary endpoints were each component of the primary endpoint. Target vessel revascularization is defined as any repeat PCI or surgery with CABG of the target lesion for ischemic symptoms and events.

### Statistical analysis

The normality of continuous variables was assessed using the Shapiro-Wilk test. Continuous data are expressed as mean ± standard deviation (SD), and skewed data are presented as medians with interquartile ranges. Unpaired Student’s *t*-test and Chi-squared test were performed to compare continuous and categorical variables, respectively. The Mann-Whitney *U*-test was performed for skewed data. Univariate and multivariate analyses with Cox proportional hazards regression were used to determine significant predictors of clinical events. Predictors that were significant in the univariate analysis were included in the multivariate analysis. Cumulative overall and event-free survival rates were computed using the Kaplan-Meier method and compared using the log-rank test. Statistical significance was set at P < 0.05. All statistical analyses were performed using R studio version 2023. 12. 1 + 402.

## Results

### Baseline Characteristics

The baseline characteristics of the 252 patients who underwent PCI with DCB are shown in Table 1. The mean age was 68.8 ± 11.9 years, and there were 204 men (81%). Hypertension, dyslipidemia, diabetes mellitus, smoking, and AF were identified in 208 (83%), 142 (56%), 116 (46%), 172 (68%), and 39 (15%), respectively. Family history of CAD, previous MI, previous PCI, previous CABG, previous stroke, CHF, PAD, AAA, COPD, and malignant tumor were identified in 27 (11%), 58 (23%), 96 (38%), 10 (4%), 32 (13%), 61 (24%), 20 (8%), 14 (6%), 18 (7%), and 36 (14%), respectively. The clinical presentations of ACS were observed as ST elevation MI (STEMI) in 30 (12%) patients, non-STEMI (NSTEMI) in 54 (21%) patients, and unstable angina in 14 (6%) patients. There were 154 (61%) patients with CCS. Three vessel disease and left main disease were identified in 35 (14%) and 11 (4%) patients, respectively. The prevalence of chronic total occlusion (CTO) was 37 (15%). Sixty patients were treated in combination with DES. The mean left ventricular ejection fraction was 54.1 ± 13.5%.

**Table 1.** Baseline clinical Characteristics.

|  | All patients<br>(n = 252) | No CKD<br>(n = 130) | CKD<br>(n = 122) | P value |
| --- | --- | --- | --- | --- |
| Age, years | 68.8 ± 11.9 | 66.8 ± 12.3 | 70.8 ± 11.2* | 0.007 |
| Males, n (%) | 204 (81) | 115 (88) | 89 (73) | 0.003 |
| Body mass index, kg / m<sup>2</sup> | 24.9 ± 4.0 | 25.4 ± 3.7 | 24.4 ± 4.2 | 0.059 |
| Hypertension, n (%) | 208 (83) | 97 (75) | 111 (91) | 0.001 |
| Dyslipidemia, n (%) | 142 (56) | 73 (56) | 69 (57) | 1.000 |
| Diabetes mellitus, n (%) | 116 (46) | 47 (36) | 69 (57) | 0.002 |
| Smoking, n (%) | 172 (68) | 97 (75) | 75 (62) | 0.035 |
| AF, n (%) | 39 (15) | 19 (15) | 20 (16) | 0.829 |
| Family history of CAD, n (%) | 27 (11) | 20 (15) | 7 (6) | 0.023 |
| Previous MI, n (%) | 58 (23) | 23 (18) | 35 (29) | 0.055 |
| Previous PCI, n (%) | 96 (38) | 41 (32) | 55 (45) | 0.037 |
| Previous CABG, n (%) | 10 (4) | 1 (1) | 9 (7) | 0.018 |
| Previous Stroke, n (%) | 32 (13) | 10 (8) | 22 (18) | 0.023 |
| Chronic heart failure, n (%) | 61 (24) | 12 (9) | 49 (40) | < 0.001 |
| PAD, n (%) | 20 (8) | 9 (7) | 11 (9) | 0.703 |
| AAA, n (%) | 14 (6) | 5 (4) | 9 (7) | 0.343 |
| COPD, n (%) | 18 (7) | 11 (9) | 7 (6) | 0.552 |
| Malignant tumor, n (%) | 36 (14) | 19 (15) | 17 (14) | 1.000 |
| Clinical presentation, n (%) |  |  |  |  |
| STEMI | 30 (12) | 19 (15) | 11 (9) | 0.239 |
| NSTEMI | 54 (21) | 31 (24) | 23 (19) | 0.417 |
| Unstable angina | 14 (6) | 10 (8) | 4 (3) | 0.210 |
| CCS | 154 (61) | 70 (54) | 84 (69) | 0.021 |
| Complexity of disease, n (%) |  |  |  |  |
| 1VD | 140 (56) | 80 (62) | 60 (49) | 0.065 |
| 2VD | 66 (26) | 32 (25) | 34 (28) | 0.657 |
| 3VD | 35 (14) | 12 (9) | 23 (19) | 0.043 |
| LMT | 11 (4) | 6 (5) | 5 (4) | 1.000 |
| CTO, n (%) | 37 (15) | 13 (10) | 24 (20) | 0.047 |
| DES-hybrid treatment, n (%) | 60 (24) | 26 (20) | 34 (28) | 0.188 |
| Echocardiographic data |  |  |  |  |
| LVEF, % | 54.1 ± 13.5 | 57.7 ± 11.7 | 50.2 ± 14.3 | < 0.001 |
| Medications, n (%) |  |  |  |  |
| Aspirin, n (%) | 230 (91) | 118 (91) | 112 (92) | 0.946 |
| P2Y<sub>12</sub> Inhibitors, n (%) | 235 (93) | 119 (92) | 116 (95) | 0.385 |
| Oral anticoagulant agents, n (%) | 43 (17) | 21 (16) | 22 (18) | 0.819 |
| Statins, n (%) | 215 (85) | 115 (89) | 100 (89) | 0.201 |
| RASi, n (%) | 182 (72) | 99 (76) | 83 (74) | 0.194 |
| β blockers, n (%) | 150 (60) | 66 (51) | 84 (69) | 0.005 |

Data are presented as mean ± S.D or % unless otherwise indicated; IQR, interquartile range; AF, atrial fibrillation ; CAD, coronary artery disease; MI, myocardial infarction; PCI, percutaneous coronary intervention; CABG, coronary artery bypass graft; CKD, chronic kidney disease; PAD, peripheral artery disease; AAA, abdominal aortic aneurysm; COPD; chronic obstructive pulmonary disease; STEMI, ST-segment elevation myocardial infarction; NSTEMI, non-STEMI; CCS, chronic coronary syndrome; VD, vessel disease; LMT, left main coronary trunk; CTO, chronic total occlusion; DES, drug eluting stent; LVEF, left ventricular ejection fraction; RASi, renin angiotensin system inhibitor.

Patients were divided into two groups according to the prevalence of CKD. Patients with CKD were significantly older and more women than those without CKD. The prevalence of hypertension, diabetes mellitus, previous PCI, previous CABG, previous stroke, and CHF were significantly higher in patients with CKD than in those without CKD. Patients with CKD had lower prevalence rates of smoking and family history of CAD. CCS, three vessel diseases, and CTO were more likely observed in patients with CKD than in those without CKD. LVEF was significantly lower in patients with CKD than in those without CKD. Patients with CKD took more β blockers than those without CKD. There were no significant differences in the other clinical factors between the two groups (Table 1).

### Association between CKD and Clinical Outcomes

During the follow-up period, there were 70 composite clinical events including 19 all-cause deaths, 12 MI, 34 target vessel revascularization, 4 stroke, and 1 bleeding.

The Kaplan-Meier analysis demonstrated that composite clinical events were higher in patients with CKD who underwent PCI with DCB than in those without CKD (log-rank test, P = 0.003) (Figure 1).

**Fig 1.**
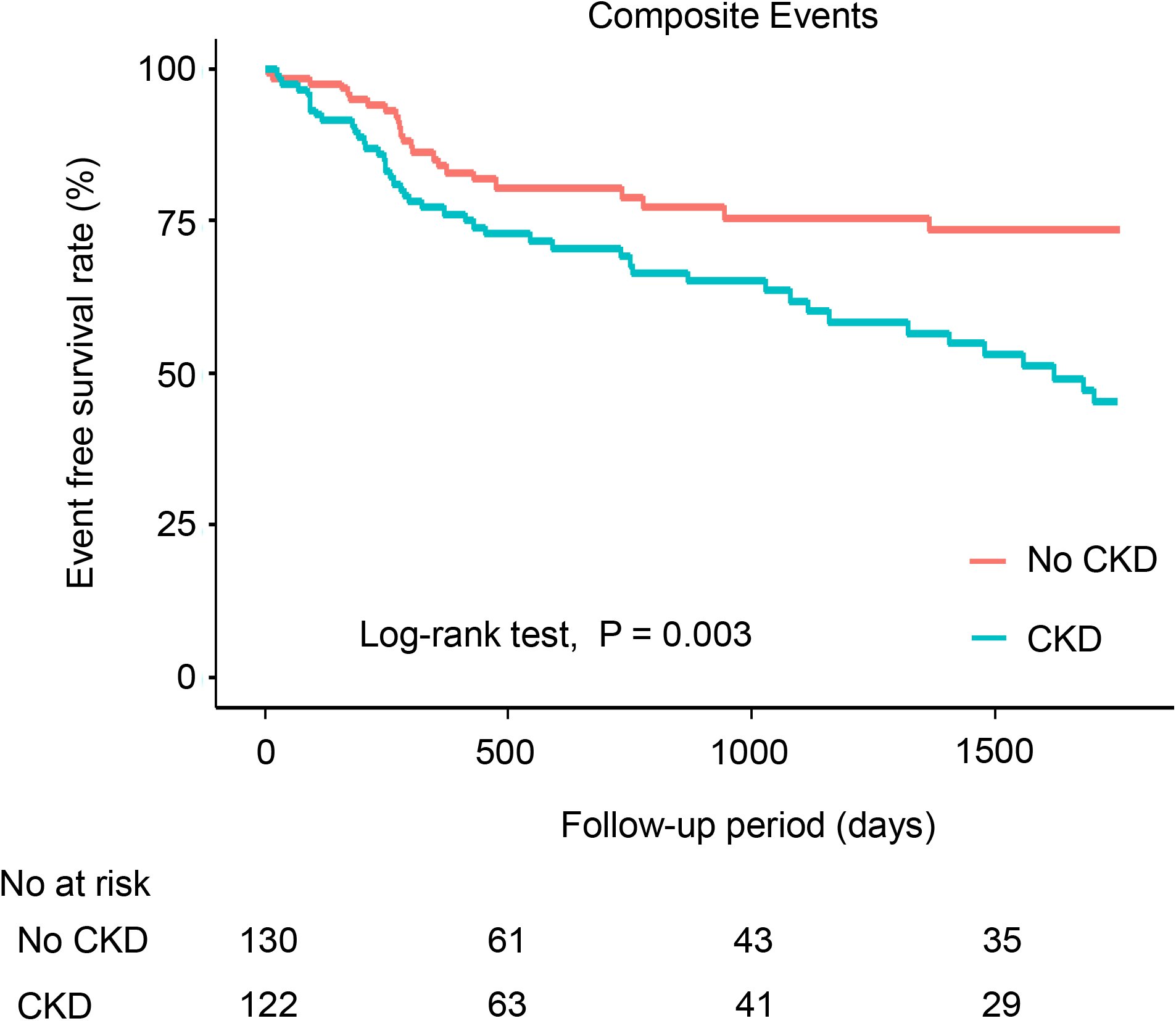
Cumulative incidence of composite events according to CKD status. Kaplan-Meier analysis demonstrated a significantly higher composite event rate in patients with CKD (log-rank test, P = 0.003). CKD, chronic kidney disease.

In the univariate Cox proportional hazard analysis, CKD and LVEF were significantly associated with the composite events. Multivariate Cox proportional hazard analysis showed that CKD was independently associated with the composite events after adjusting for LVEF (hazard ratios, 1.920; 95% confidence intervals, 1.154-3.197; P = 0.012) (Table 2).

**Table 2.** Univariate and multivariate Cox proportional hazard analyses for composite events.

| Variables | HR | 95% CI | P value |
| --- | --- | --- | --- |
| Univariate analysis |  |  |  |
| Age | 1.006 | 0.986-1.025 | 0.580 |
| Male | 0.975 | 0.551-1.727 | 0.932 |
| Body mass index | 0.941 | 0.883-1.004 | 0.065 |
| Hypertension | 1.338 | 0.664-2.698 | 0.415 |
| Dyslipidemia | 1.006 | 0.625-1.620 | 0.980 |
| Diabetes mellitus | 1.523 | 0.950-2.437 | 0.081 |
| smoking | 1.200 | 0.728-1.979 | 0.475 |
| AF | 1.070 | 0.547-2.094 | 0.843 |
| Family history of CAD | 0.512 | 0.186-1.404 | 0.193 |
| Previous MI | 1.231 | 0.726-2.088 | 0.441 |
| Previous PCI | 1.562 | 0.977-2.499 | 0.063 |
| Previous CABG | 2.230 | 0.892-5.574 | 0.086 |
| Previous Stroke | 1.120 | 0.556-2.257 | 0.750 |
| Chronic heart failure | 1.394 | 0.822-2.365 | 0.217 |
| CKD | 2.076 | 1.267-3.402 | 0.004 |
| PAD | 1.895 | 0.900-3.992 | 0.093 |
| AAA | 1.337 | 0.537-3.328 | 0.532 |
| COPD | 1.394 | 0.559-3.474 | 0.476 |
| Malignant tumor | 1.175 | 0.600-2.302 | 0.638 |
| STEMI | 1.291 | 0.678-2.458 | 0.438 |
| NSTEMI | 0.937 | 0.521-1.684 | 0.827 |
| Unstable angina | 0.888 | 0.279-2.825 | 0.840 |
| CCS | 0.938 | 0.581-1.515 | 0.795 |
| 1VD | 0.775 | 0.484-1.241 | 0.288 |
| 2VD | 1.084 | 0.627-1.873 | 0.772 |
| 3VD | 1.200 | 0.630-2.284 | 0.580 |
| LMT | 1.831 | 0.667-5.028 | 0.241 |
| CTO | 1.054 | 0.540-2.056 | 0.877 |
| DES-hybrid treatment | 0.663 | 0.363-1.211 | 0.181 |
| LVEF | 0.983 | 0.967-0.999 | 0.042 |
| Aspirin | 4.186 | 0.581-30.180 | 0.155 |
| P2Y<sub>12</sub> Inhibitors | 0.473 | 0.216-1.037 | 0.062 |
| Oral anticoagulant agents | 1.040 | 0.545-1.983 | 0.905 |
| Statins | 0.761 | 0.440-1.318 | 0.330 |
| RASi | 1.161 | 0.691-1.951 | 0.574 |
| β blockers | 0.961 | 0.764-1.208 | 0.732 |
| Multivariate analysis |  |  |  |
| CKD | 1.920 | 1.154-3.197 | 0.012 |
| LVEF | 0.989 | 0.972-1.006 | 0.198 |

HR, hazard ratio; CI, confidence interval; AF, atrial fibrillation ; CAD, coronary artery disease; MI, myocardial infarction; PCI, percutaneous coronary intervention; CABG, coronary artery bypass graft; CKD, chronic kidney disease; PAD, peripheral artery disease; AAA, abdominal aortic aneurysm; COPD; chronic obstructive pulmonary disease; STEMI, ST-segment elevation myocardial infarction; NSTEMI, non-STEMI; CCS, chronic coronary syndrome; VD, vessel disease; LMT, left main coronary trunk; CTO, chronic total occlusion; DES, drug eluting stent; LVEF, left ventricular ejection fraction; RASi, renin angiotensin system inhibitor.

The higher composite event risk was mainly driven by an increased risk of all-cause death (Figure 2 and Table 3).

**Fig 2.**
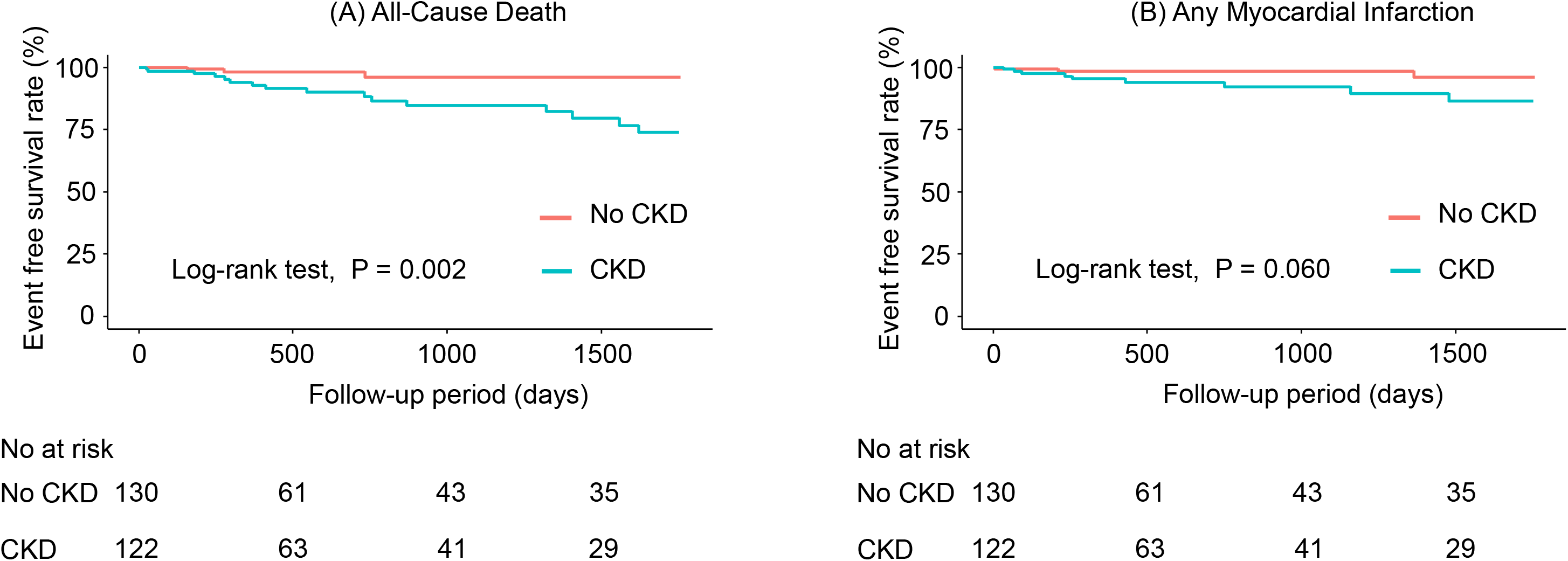
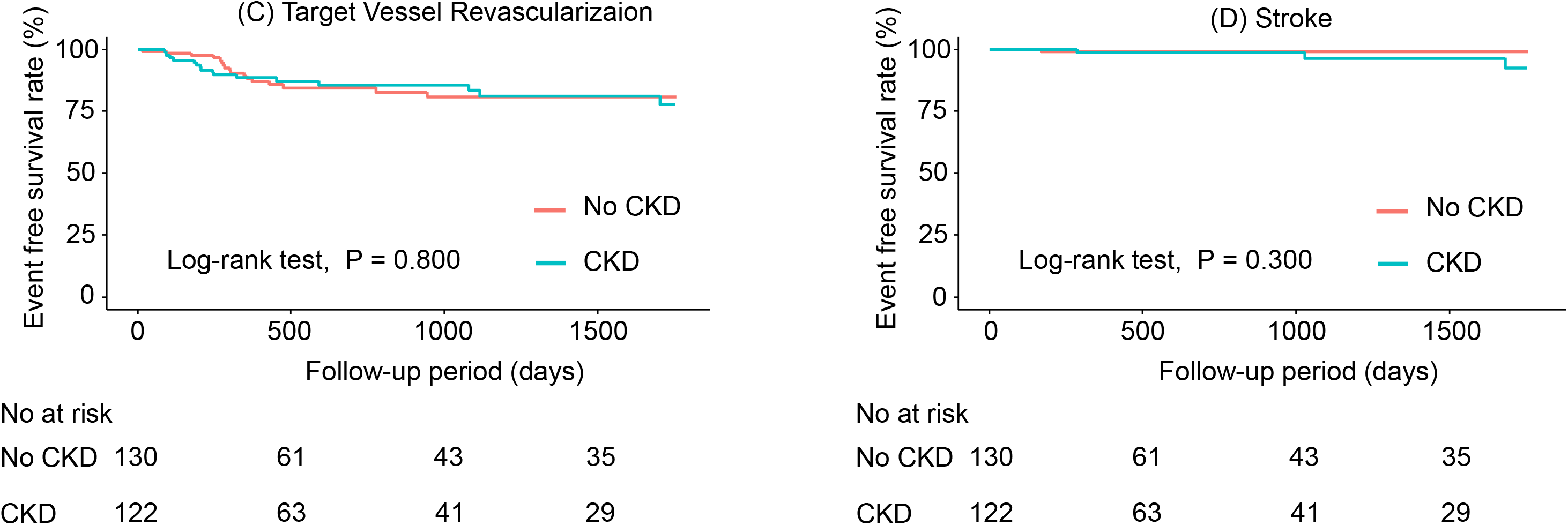
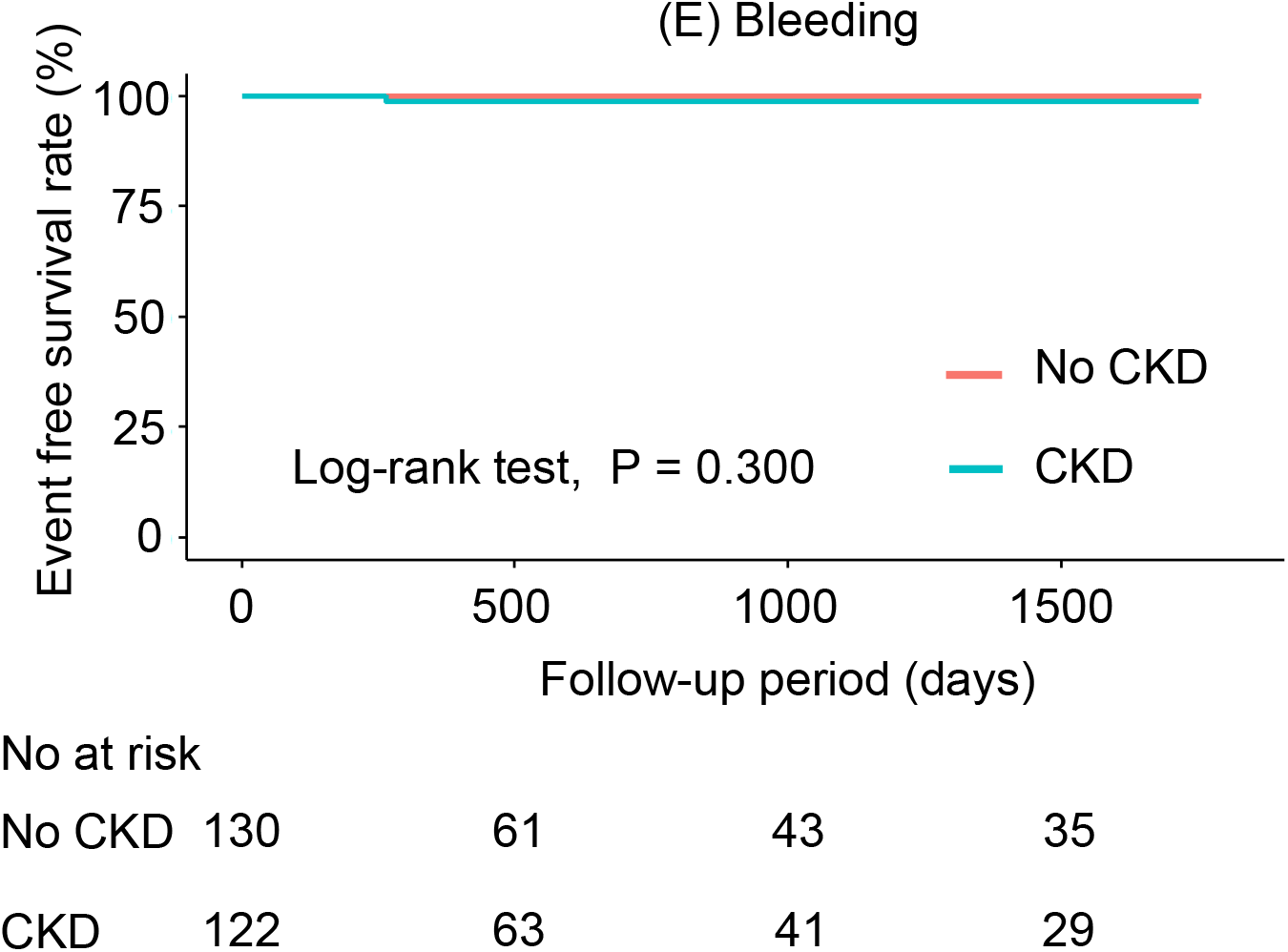
Cumulative incidence of (A) all-cause death, (B) any myocardial infarction, (C) target vessel revascularization, (D) stroke, and (E) bleeding according to CKD status. Kaplan-Meier analysis demonstrated a significantly higher rate of all-cause death in patients with CKD (log-rank test, P = 0.002). No differences in term of myocardial infarction, target vessel revascularization, strike, and bleeding were observed. CKD, chronic kidney disease.

**Table 3.** Comparison of clinical outcomes according to CKD status.

|  | No CKD<br>(n = 130) | CKD<br>(n = 122) | HR<br>(95% CI) | P value | Adjusted HR<br>(95% CI) | P value |
| --- | --- | --- | --- | --- | --- | --- |
| All-Cause Death | 3 (2.3%) | 16 (13.1%) | 5.689<br>(1.656-19.540) | 0.006 | 5.224<br>(1.492-18.291) | 0.010 |
| Any Myocardial Infarction | 3 (2.3%) | 9 (7.4%) | 3.282<br>(0.889-12.140) | 0.075 | 2.800<br>(0.732-10.721) | 0.133 |
| Target Vessel Revascularization | 17 (13.1%) | 17 (13.9%) | 1.081<br>(0.552-2.119) | 0.820 | 1.049<br>(0.521-2.110) | 0.893 |
| Stroke | 1 (0.8%) | 3 (2.5%) | 3.390<br>(0.352-32.620) | 0.290 | 2.684<br>(0.267-26.991) | 0.402 |
| Bleeding | 0 (0%) | 1 (0.8%) | NA | NA | NA | NA |

CKD, chronic kidney disease; HR, hazard ratio; CI, confidence interval; NA, not applicable.

Further, the patients were divided into the five groups according to the CKD stages classified by eGFR (G1, ≥ 90 mL/min/1.73m^2^; G2, 60 – 89 mL/min/1.73m^2^; G3, 30 – 59 mL/min/1.73m^2^; G4, 15 – 29 mL/min/1.73m^2^; G5, < 15 mL/min/1.73m^2^) ^20^. Kaplan-Meier analysis revealed that composite events were increased with advancing CKD stages (log-rank test, P < 0.0001) (Supplemental Figure 1).

## Discussion

In the present study, (1) Kaplan-Meier analysis demonstrated a significantly higher event rate in patients with CKD than in those without; and (2) multivariate Cox hazard analysis revealed that CKD was significantly associated with poor clinical outcomes in CAD patients who underwent PCI with DCB, driven by higher all-cause mortality.

Recently, outcomes of PCI have been dramatically improved since various technologies of interventional cardiology have been developed. Especially, the evolution of DES and procedural techniques, and use of intravascular imaging have made a great contribution to improvement of outcomes of PCI ^22^. Furthermore, optimal short duration of dual antiplatelet therapy (DAPT) after PCI has succeeded in favorable prognosis ^23^. However, there are still residual risks of PCI such as stent-related adverse events in late phase after PCI even in the era of contemporary DES. A recent study demonstrated that very-late stent-related events occurred in 1 to 5 years after PCI regardless of the type of stents, and the rate of events was about 2% per year with no plaque evident ^24^.

Contemporary stent technologies have not succeeded in conquest of this ongoing very-late stent-related event risks ^24^. Stent under-expansion, malaposition, uncovered stent strut, hypersensitivity reactions, stent fracture, and neoatherosclerosis have been reportedly associated with very-late stent-related events ^25^. Therefore, a treatment strategy that leaves nothing in coronary artery has been needed.

DCB is a novel technology of PCI and a well established treatment option for contemporary PCI. DCB directly releases antiproliferative drugs into the vessel wall of coronary artery, which leads to the reduction of the untoward effects associated with polymer-based stent technologies ^26^. In the clinical setting of coronary in-stent restenosis, treatment with DCB significantly lowered the incidence of adverse events and recurrent in-stent restenosis compared to drug uncoated balloon ^1^. For the treatment of small native coronary arteries, DCB also showed non-inferior event rates compared to DES with respect to major adverse cardiovascular events (MACE) for up to 3 years ^2, 3^. Recently, DCB treatment in combination with DES in CAD patients with multivessel diseases significantly reduced MACE at 2 years ^4^. Furthermore, recent study have demonstrated that DCB has a potential to improve the cardiovascular outcomes compared to DES even in de novo large coronary arteries ^5^. Thus, DCB has various favorable impacts on clinical outcomes in CAD patients who underwent PCI in the era of contemporary DES. However, the prognostic factors of DCB on clinical outcomes in CAD patients who underwent PCI have not been fully described. Further, the favorable clinical evidences about DCB are almost in the setting of comparison between DCB and DES as described above ^1, 2, 3, 4, 5^.

In the present study, we demonstrated that CKD was associated with poor clinical outcomes in CAD patients who underwent PCI with DCB. The prevalence rate of CKD in patients with CAD who underwent PCI has been increasing due to the aging population ^8^. CAD patients with CKD showed worse clinical outcomes than those with preserved renal function ^11^. A recent study demonstrated that angiographic complete revascularization using DES led to favorable prognosis in CAD patients with CKD ^27^. However, there are few data about the prognostic impact of CKD on PCI with DCB. The subgroup analyses of the BASKET-SMALL 2 demonstrated that the clinical outcomes were not different between DCB and DES in patients of CKD ^2^. The present study revealed the association between the presence of CKD and unfavorable prognosis in CAD patients who underwent PCI with DCB.

The pathophysiological role of CKD on the poor prognosis of CAD patients who underwent PCI with DCB has not been fully addressed. In the present study, the proportion of severe CAD status such as 3 vessel disease and CTO was significantly higher in patients with CKD than in those without. The presence of CKD was reported to be associated with the risk of increased coronary lesion complexity in patients with CAD ^10^. In CAD patients who underwent PCI with DCB, the presence of CKD might affect the progression of CAD, which leads to poor clinical outcomes.

In the present study, the poor prognosis in CKD patients was mainly driven by an increased risk of all-cause death. The previous study of PCI with DES, the all-cause mortality in CKD patients was about 10% ^27^, which was consistent with the results of this study. The incident any MI was also higher in patients with CAD although it did not reach statistical significance. This result suggested that the presence of CKD may accelerate atherosclerotic lesion progression and leads to unfavorable prognosis after PCI with DCB.

### Study limitations

This study has several limitations. First, this was a single-center study with a relatively small sample size. Large study population is needed to further clarify prognostic factor in patients with CAD who underwent PCI with DCB, although the sample sizes of recent studies about DCB were relatively small ^45^. Second, there were not detailed data of procedure of PCI such as length and diameter of DCBs, length of culprit lesion, and contrast volume because the present study did not focus on PCI procedure. Since the presence of CKD has been reported to be associated with the progression of atherosclerosis ^10^, the PCI procedure might be different between 2 groups with and without CKD, which might affect clinical outcomes in the present study. Third, the data of complete revascularization was missing in the present study. The previous study demonstrated that complete revascularization was associated with favorable prognosis in CAD patients with CKD who underwent PCI with DES ^27^. There is a possibility that the presence of complete revascularization might affect the results in the present study. Forth, we did not analyze the data of intracoronary imaging, although the usage rate of intracoronary imaging devices was 98% in the present study. The detailed data of intracoronary imaging devices might provide important information of the role of CKD on unfavorable prognosis in CAD patients who underwent PCI with DCB. Fifth, the pathological mechanism of CKD in patients who underwent PCI with DCB was unclear. Vascular calcification and inflammation were reportedly associated with vascular disease in CKD patients ^28^. Further studies are required to clarify the extent of these limitations.

## Conclusions

CKD was associated with unfavorable outcomes in patients with CAD who underwent PCI with DCB. The presence of CKD provides useful information for the risk stratification of CAD patients who underwent PCI with DCB.

## Data Availability

The data underlying this article are available in the article and in its supplemental materials.

## Disclosures

The authors declare no conflicts of interest.

## Acknowledgments

The authors thank Editage (www.editage.jp) for English language review.

## Notes

### Competing Interest Statement

The authors have declared no competing interest.

### Author Declarations

The study was approved by the Institutional Ethics Committee of Yamagata University School of Medicine (Yamagata University, 2020-344).

