## Supplementary figures and images for "Impact of Chronic Kidney Disease on Clinical Outcomes after Percutaneous Coronary Intervention with Drug-Coated Balloons"

### Supplemental Figure

Supplemental Figure 1.

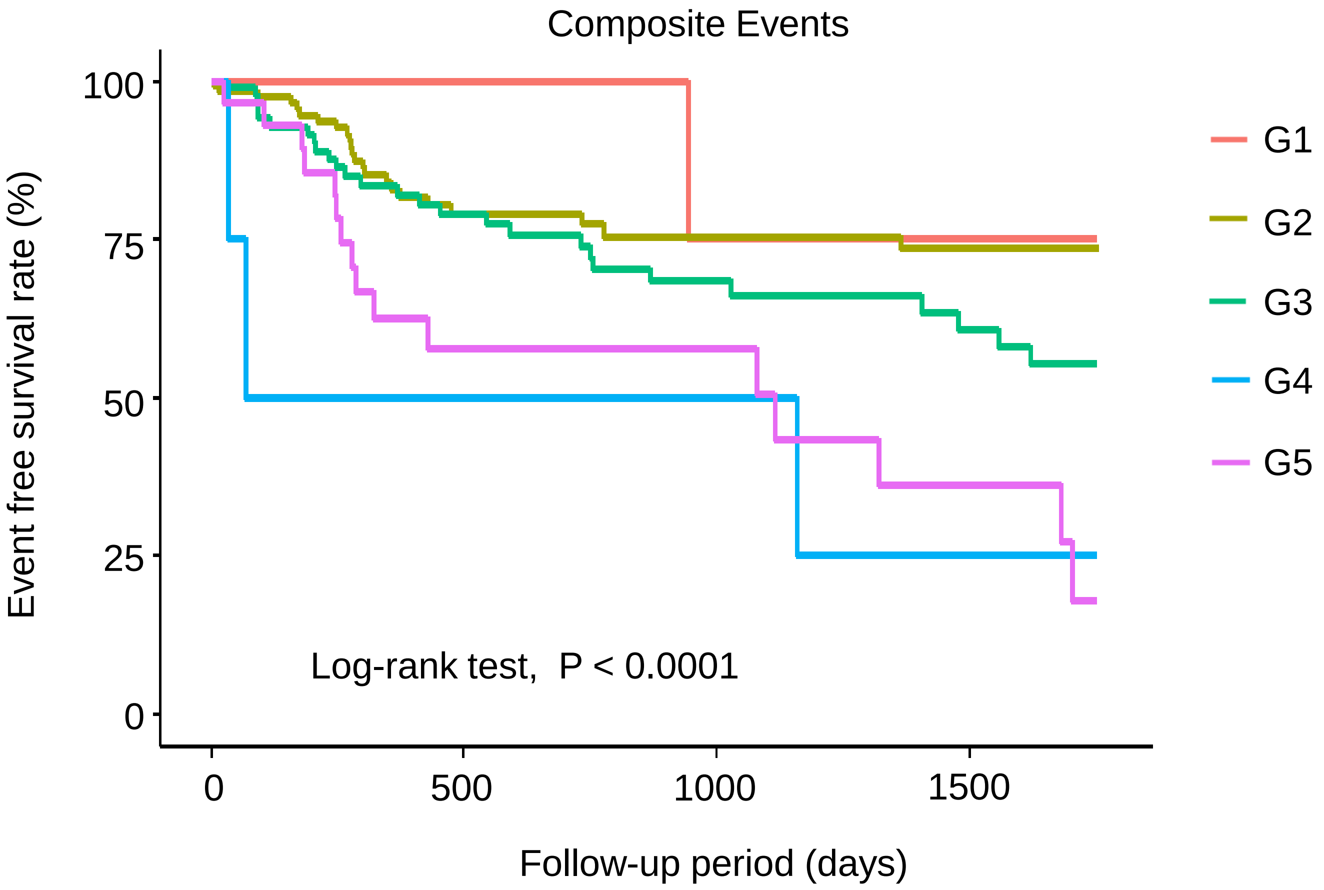


No at risk

| | | | | |
|---|---|---|---|---|
| G1 | 11 | 6 | 3 | 3 |
| G2 | 119 | 55 | 40 | 32 |
| G3 | 88 | 49 | 31 | 23 |
| G4 | 4 | 2 | 2 | 1 |
| G5 | 30 | 12 | 8 | 5 |
